# Neuropsychological stratification of Chronic Obstructive Pulmonary Disease: building affect into clinically relevant neuro-biomarkers of breathlessness

**DOI:** 10.1101/19006684

**Authors:** Sarah L. Finnegan, Olivia K. Faull, Catherine J. Harmer, Mari Herigstad, Najib M. Rahman, Andrea Reinecke, Kyle T.S. Pattinson

## Abstract

**Background:** Chronic breathlessness profoundly affects quality of life for its sufferers. Often, reported breathlessness is inconsistent with airway pathophysiology and objective disease markers. While a mechanistic understanding of this discordance has thus far remained elusive, factors such as mood, attention and expectation have all been implicated as important perceptual modulators. Therefore, here we have developed a model capable of exploring these relationships aiding patient stratification and revealing clinically-relevant neuro-biomarkers.

**Methods:** A cohort of 100 participants with mild-to-moderate chronic obstructive pulmonary disease (COPD) underwent a comprehensive assessment that included functional brain imaging while viewing and rating breathlessness-related word cues, self-report questionnaires and clinical measures.

**Results:** Using an exploratory factor analysis across psychological and physiological measures, we identified two distinctive neuropsychological behavioural profiles that differed across four key factors corresponding to mood, symptom burden, and two capability measures. These profiles stratified participants into high and low symptom groups, which did not differ in spirometry values. The low symptom load group demonstrated greater FMRI activity to breathlessness-related word cues in the anterior insula.

**Conclusions:** Our findings reveal two clear groups of individuals within our COPD cohort, divided by behavioural rather than clinical factors. Furthermore, indices of depression, anxiety, vigilance and perceived capability were linked to differences in brain activity within key regions thought to be involved in monitoring bodily sensations (interoception). These findings demonstrate the complex relationship between affect and interoceptive processing, providing the foundations for the development of targeted treatment programmes that harness clinical and symptom-relevant biomarkers.

## Introduction

For the millions of people living with chronic obstructive pulmonary disease (COPD), asthma, heart failure or cancer, breathlessness is a major source of suffering, extending pervasively into people’s lives. COPD describes a collection of lung conditions including bronchitis and emphysema, 85% of which arise as a result of long-term cigarette smoking. Despite the substantial personal, social and economic impacts of breathlessness resulting from COPD alone (4.5% of people over 40 have been diagnosed with COPD according to the British lung foundation and in 2015 5.6% of the worldwide deaths were attributed to COPD [1]), mechanistic understandings of symptom variability and the discordance between pathophysiology and subjective perceptions remain elusive.

While traditional thinking has viewed breathlessness as a symptom arising directly from lung damage or cardiopulmonary stress, recent work in breathlessness has highlighted the influence of factors such as mood and attention on subjective perceptions [2]. Furthermore, 40% of people with COPD are thought to experience clinically relevant anxiety and depression, compared to <10% of the general population [2]. In addition to these factors, previous experiences, symptom expectation and the ability to correctly monitor our internal bodily state (also known as interoception [3, 4]) also appear to play a fundamental role in perceptual experiences, as outlined by the “Bayesian Brain” hypothesis.

In the Bayesian Brain model, the brain weighs prior expectations against noisy ascending sensory inputs to determine sensory perception. This perceptual weighting is thought to be further modulated by emotions and attention [5]. Such theories are beginning to gain traction and formalising these in terms of their neural correlates may yet offer a fresh perspective on the challenges of understanding breathlessness [6, 7].

Thus, breathlessness arises not only from current patho-physiological status of the lungs and airways, but also depends on the brain’s interoceptive networks – those networks associated with interpreting incoming signals from the body. The production of breathlessness perceptions arise from a complex product of previous experiences, the interpretation of internal sensory signals and factors such as emotional status. A model incorporating this interplay of the behavioural and neuro-psychological networks may facilitate stratification of patient groups based on brain-related metrics, offer explanations for symptom discordance and heterogeneity and in turn lead to the development of targeted treatments within these groups.

Here, we have investigated this relationship by building on our previous work linking brain activity to breathlessness-related word cues [8, 9] and drawing upon the big-data techniques of unsupervised machine learning and dimensionality reduction. We applied unsupervised machine learning algorithms to a large multi-modal questionnaire, behavioural and physiological dataset and identified the best combination of measures for describing the breathlessness experience across individuals. Using dimensionality reduction, we then reduced the measures down to their underlying factors and used these to stratify the population of individuals with COPD into subgroups with distinct neuropsychological profiles. Finally we used this information to probe potential differences in neural processing of breathlessness expectations between these symptom-based groups.

## Methods and Materials

A brief overview of the study methodology is presented here. Full details can be found within supplementary materials.

### Participants

100 participants (36 female, mean age 69 years (49-84 years)) with mild to moderate COPD (according to GOLD [Global Initiative for Chronic Obstructive Lung Disease] standards) were recruited to this study immediately prior to their enrolment in a National Health Service prescribed course of pulmonary rehabilitation. From this population, 91 participants completed the magnetic resonance imaging (MRI) component of the study (Supplement Figure 1). Written informed consent was obtained from all participants prior to the start of the study. Study approval was granted by South Central Oxford REC B (Ref: 118784).

### Behavioural Measures

#### Self-report questionnaires

We selected a comprehensive set of questionnaires designed to probe the experience of living with COPD, focusing particularly on the potential influence of emotional and behavioural measures.

The following self-report questionnaires were completed and scored according to their respective manuals: Dyspnoea-12 (D12) Questionnaire [10], Centre for Epidemiologic Studies Depression Scale (CES-D) [11], Trait Anxiety Inventory (TRAIT) [12], Fatigue Severity Scale [13], St George’s Respiratory Questionnaire (SGRQ) [14], Medical Research Council (MRC) breathlessness scale [15], Mobility Inventory (MI) [16], Pittsburgh Sleep Quality Index [17], Anxiety Sensitivity Index Questionnaire (ASI) [18], Catastrophic Thinking Scale in Asthma [19], Pain Awareness and Vigilance Scale [20].

#### Emotional Stroop

Participants completed an emotional Stroop task, featuring breathlessness-related words vs control neutral words matched for length and frequency (modified from: *Reinecke et al*., *2011*). Participants were required to read aloud the colour of the word and the reaction time to respond to emotional vs neutral words was computed. Due to technical issues, Stroop data were collected from a subgroup of 66 participants.

#### Physiological Measures

Spirometry and two Modified Shuttle Walk Tests (MSWT) were collected using standard practices. Participant height and weight was recorded at each session.

### MRI Measures

#### Image acquisition

Imaging was carried out using a 3T MAGNETOM Trio, A Tim System (Siemens Healthcare GmbH) using a 12-channel head coil. A T1-weighted (MPRAGE) structural scan (voxel size: 1 × 1 × 1 mm) was collected and used for registration purposes. A T2*-weighted, gradient echo planar image (EPI) scan sequence (voxel size: 3 × 3 × 3 mm) was used to collect functional imaging data during the word cue task. Field map scans of the B0 field were obtained to aid the distortion correction of the functional scans

#### Word cue task

To probe breathlessness-related expectation we examined the activity of brain regions responding to breathlessness-related word cues and fixed string non-words, and correlated this to corresponding visual analogue ratings of anxiety and breathlessness [22]. Ratings were recorded throughout the task and word presentation order was pseudo-randomized for each session.

#### Faces task

In the second functional scan, participants were shown human faces for 500ms (drawn from the set described by [23] in blocks of 30 seconds, with fearful or happy expressions. Participants were asked to identify whether the faces were male or female. A fixation cross condition was interspersed for 30 seconds between the happy/fearful blocks of faces. Reaction time and accuracy were recorded throughout the task.

## Analysis

### Behavioural Analysis – factor identification

Full correlation matrices were calculated for (z-scored) behavioural (questionnaires) and physiological scores (spirometry, demographics and MSWT measures) (Table 1), using MATLAB 2017b (Mathworks, Natick, MA). For this analysis, only data from participants (n=91) who completed both the behavioural and MRI parts of the study were utilised. The structure of the correlation matrices was examined by applying a hierarchical cluster model to the data (Figure 1). To formalize the relationships observed as a result of applying the hierarchical cluster model to the data, an Exploratory Factor Analysis (EFA) was conducted using parallel analysis, oblique rotation and a maximum likelihood estimation. Model selection criteria included loading variables above 0.4 with no cross loading or freestanding variables, and significant X^2^/df ratio with Tucker-Lewis Index (TL-index) close to 1 and Root Mean Square Error of Approximation (RMSEA) < 0.06. Models were fit using Lavaan version 0.6-1 (Rosseel, 2012) in R version 3.2.1 (R Core Team).

**Figure 1.**
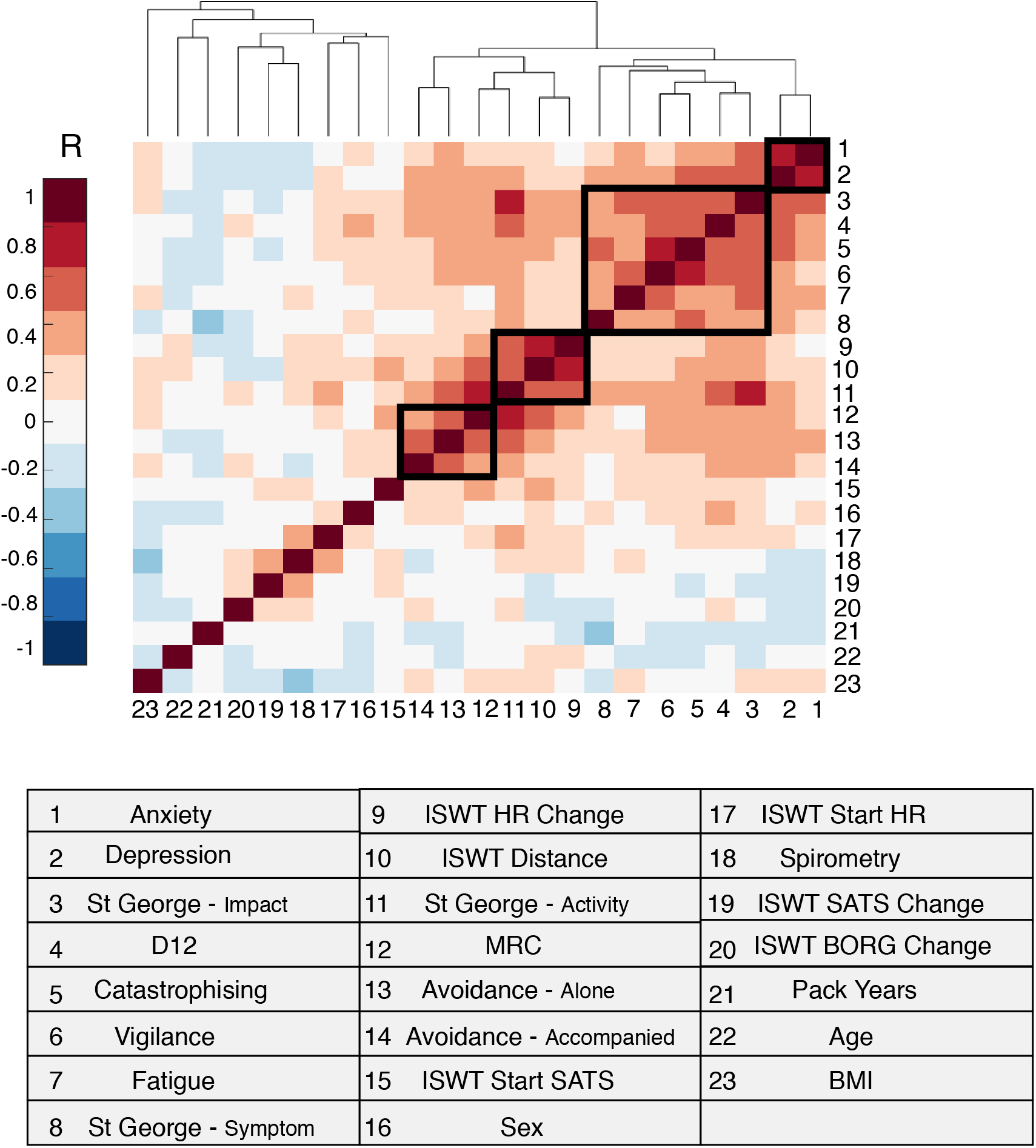
Clustergram: a correlation matrix of measured behavioural and physical variables, where strength of the correlation is measured as a Pearsons’ R-value. Variables are reordered such that more closely related measures are placed proximal to each other. The relationship between groups of measures is demonstrated by the height of the dendrogram branches and distance between neighbouring branches (in arbitrary units). Clusters identified by the EFA as significant are highlighted by black boxes.

**Table 1.** Demographic information (N=100)

|  |  |
| --- | --- |
| Age (years/range) | **69/(49-84)** |
| BMI kg.m-2 ± SD | **28.0±6.2** |
| Smoking pack-years ± SD | **36.7±26.0** |
| Resting SpO<sub>2</sub>% ± SD | **94.1±3.3** |
| Resting heart rate beats.min-1 ± SD | **80.8±13.6** |
| FEV1/FVC ± SD | **0.55±0.15** |
| FEV1 % Predicted ± SD | **58±21** |
| MRC (IQR) | **3 (1)** |

**Table 2.** A list of all measures included in the hierarchical cluster model

|  |  |  |
| --- | --- | --- |
| **1. Anxiety** | **9. MSWT HR Change** | **17. MSWT Start HR** |
| **2. Depression** | **10. MSWT Distance** | **18. Spirometry** |
| **3. St George – Impact** | **11. St George – Activity** | **19. MSWT SATS Change** |
| **4. D12** | **12. MRC** | **20. MSWT BORG Change** |
| **5. Catastrophising** | **13. Avoidance – Alone** | **21. Pack Years** |
| **6. Vigilance** | **14. Avoidance - Accompanied** | **22. Age** |
| **7. Fatigue** | **15. MSWT Start SATS** | **23. BMI** |
| **8. St George - Symptom** | **16. Sex** |  |

**Table 3.**
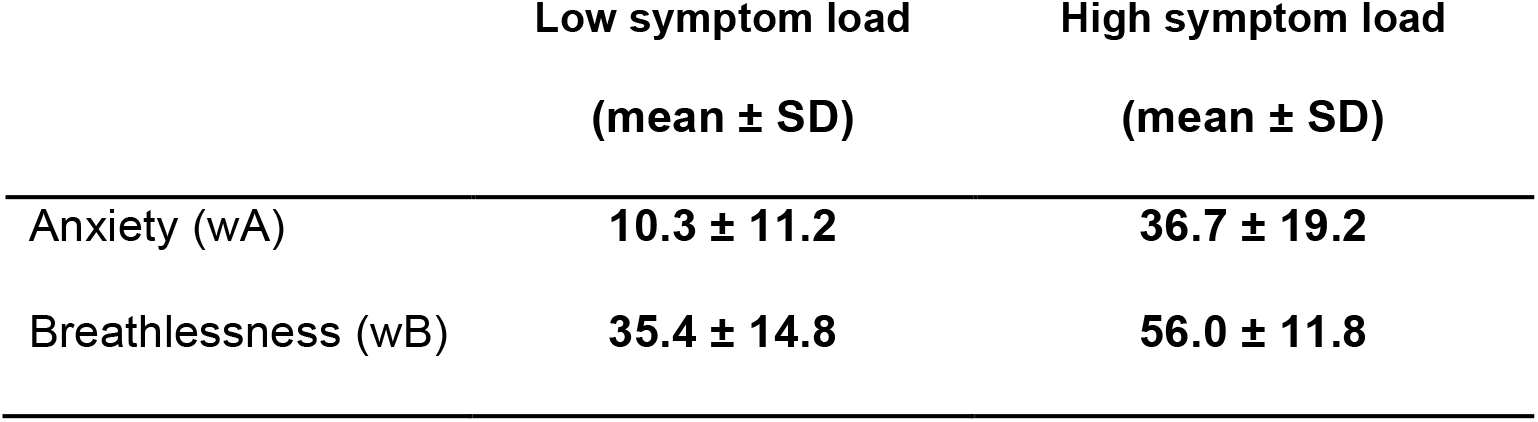
Group word cue anxiety and breathlessness scores

| | Low symptom load<br>(mean ± SD) | High symptom load<br>(mean ± SD) |
| --- | --- | --- |
| Anxiety (wA) | 10.3 ± 11.2 | 36.7 ± 19.2 |
| Breathlessness (wB) | 35.4 ± 14.8 | 56.0 ± 11.8 |

### Behavioural Analysis – participant stratification

A principal component analysis was performed across the measures within each factor, identified via the EFA in the factor identification process, reducing data-dimensionality. This created one composite score per participant for each factor. This information was input into the hierarchical cluster algorithm, alongside anonymized subject identities. A scree plot was used to identify the most statistically distinct groupings of participants (Supplementary Figure 3).

### Stroop

Stroop scores were calculated for each individual by subtracting the mean reaction time in response to neutral words from that of the social-threat words and breathlessness-threat words. This was carried out separately for the masked and unmasked conditions. Repeated measures ANOVAs were carried out separately for word type presented under masked or unmasked conditions. Labels, corresponding to high and low symptom load, assigned during the participant stratification process, were then applied to this group. Repeated measures ANOVAs were carried out separately for word type presented under masked or unmasked conditions using group identity as the predictor variable.

### Imaging Analysis

Image processing was carried out using the Oxford Centre for Functional Magnetic Resonance Imaging of Brain Software Library (FMRIB, Oxford, UK; FSL version 5.0.8; https://ww.fmrib.ox.ac.uk/fsl/), MATLAB (R2017b) and associated custom scripts.

### Functional MRI Analysis

MRI processing was performed using FEAT (FMRI Expert Analysis Tool, within the FSL package).

#### Word task lower-level analysis

At the individual subject level, a general linear model (GLM) was created with explanatory variables (EVs) for word or non-word presentation, and two de-meaned EVs modeling the reported breathlessness and anxiety response to the word cues. An additional explanatory noise variable was included to model the period during which the participant responded using the VAS. In addition to the mean contrasts for each of the EVs, differential contrasts were also created for activity in response to breathlessness-related words greater than that for non-words and for non-words greater than that for words.

#### Faces task lower-level analysis

At the individual subject level, a GLM was created with EVs for stimulus presentation periods of happy and fearful faces, along with the associated (de-meaned) reaction times. Two additional EVs were created to model participant (de-meaned) accuracy in identifying whether the presented faces were male or female. In addition to the mean contrasts for each of the EVs, differential contrasts were also created for activity in response to fearful faces greater than that for happy faces.

#### Group level analysis

Mean voxelwise difference in activity were calculated for the words > non-words, non-words > words and fearful faces > happy faces, happy faces > fearful faces contrasts between groups of participants (corresponding to high and low symptom load) identified within the hierarchical cluster model at the participant stratification stage. Demeaned age and sex values were modeled as regressors of no interest. Significance testing was performed using FSL’s Randomize tool [24] – which carries out rigorous permutation testing, with threshold free cluster enhancement (TFCE) at p<0.05. Based on *a priori* hypotheses a region of interest approach, examining differential activity specifically within the amygdala (including 10mm radius spherical masks of a left amygdala region (-14/-6/-8) and its right hemisphere counterpart) was also taken.

## Results

Of the 23 variables entered into the hierarchical cluster model (Figure 1), 14 were found to significantly contribute to a description of the variance within the COPD population. Significant variables included: Anxiety, Depression; St George – Impact, D12, Catastrophising, Vigilance, Fatigue, St George – Symptom, MSWT HR change, MSWT Distance, St George – Activity, MRC, Avoidance – Alone and MRC Avoidance - Accompanied. These groups of measures are demonstrated in Figure 1.

The final 4 factor model (shown overlaid onto Figure 1) was validated after testing for models of 2, 3 and 4 factors, (X^2^ = 65.85, df = 41, p<0.008; TLI=0.9; RMSEA=0.05). The factor diagram shown in Figure 2 shows how 4 latent factors emerged from the 14 variables. Factor 1 is made up of vigilance, catastrophising, fatigue, St George – Symptoms, St George – Impact and D12. Factor 2 was made up of MSWT Distance, MSWT HR change and St George – Activity. Factor 3 is composed of two of the Avoidance sub-scales and MRC scale. Finally Factor 4 consists depression and trait-anxiety). The covariance between factors is illustrated by the curved lines in Figure 2, with Factors 1 and 4 demonstrating the strongest covariance.

**Figure 2.**
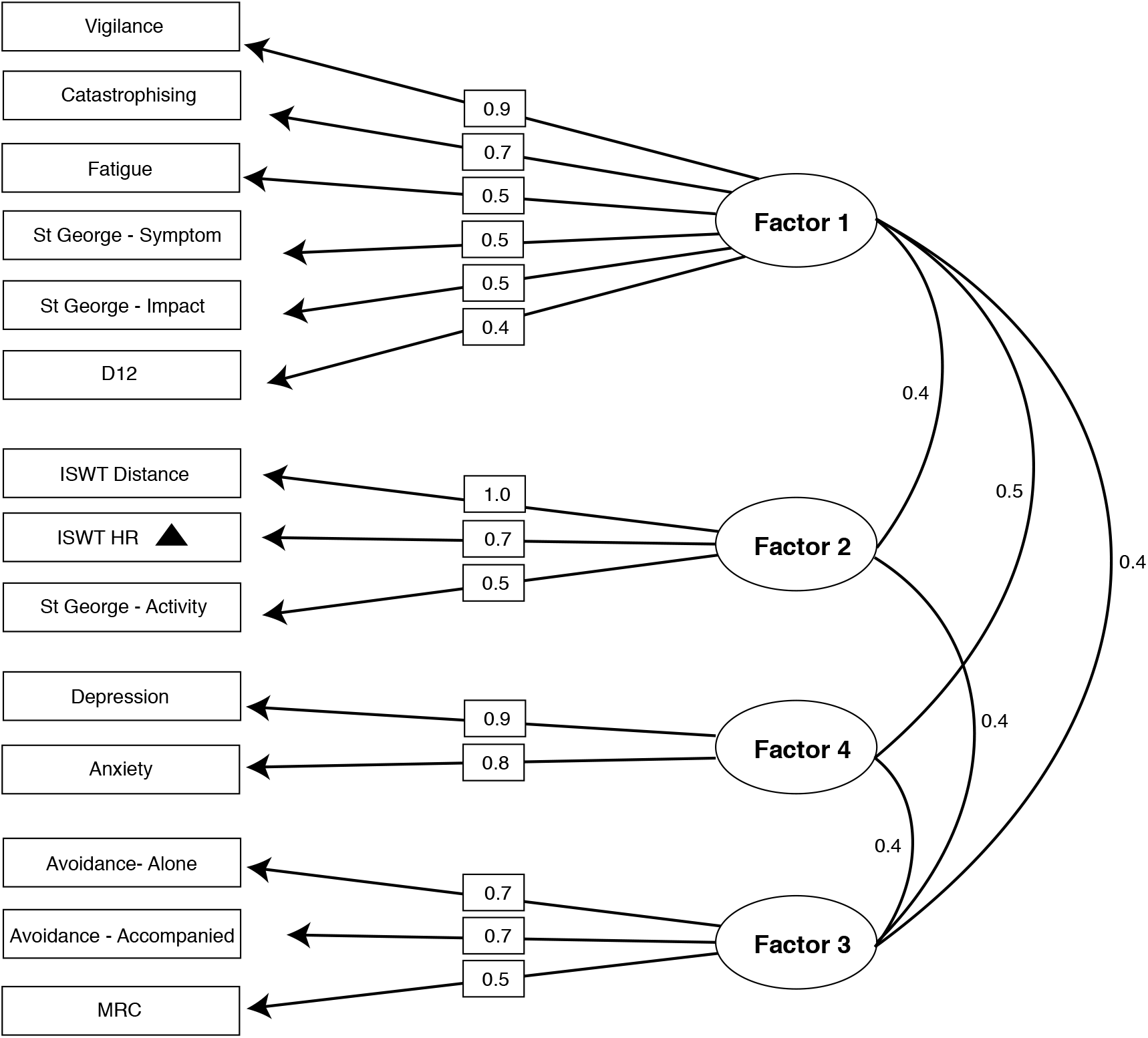
Factor Loadings: The relationship between each variable (rectangles) and its parent factor (ellipses) is shown here. Loadings (straight lines) can be interpreted as correlation coefficients. The covariance between factors (curved lines) is also shown.

The factors identified by EFA model fitting were then used to stratify the patient population via their composite scores on these 4 major factors in a hierarchical cluster model. A scree plot (Supplementary Figure 3) confirmed that a 2-group solution was the most distinct. The groupings of participants seems to correspond to high symptom load and low symptom load across the four factors - with significant paired distance across the two groups between each factor at p<0.001. This difference was not driven by differences in spirometry scores (p=0.32).

### MRI results

#### Mean group differences - word task

Robust significant group differences were found in response to the breathlessness-related word cues compared to non-words in the anterior insula (Figure 4) when using non-parametric permutation testing alongside stringent threshold-free cluster enhancement. The BOLD response within the low symptom group was found to be higher in this key region in response to breathlessness words versus non-words. No significant difference was observed within the small volume correction analysis of the amygdala. Further, more exploratory analysis is presented in Supplementary Figure 6. Participant ratings of the breathlessness-related words were found to be significantly higher in the high symptom load (p<0.01) compared to the low symptom load group for both anxiety (wA) (high load 36.7 ± 19.2, low load 10.3 ± 11.2) and breathlessness (wB) (high load 56.0 ± 11.8, low load 35.4 ± 14.8), measures that were not used in the classification of the participants into groups.

**Figure 3.**
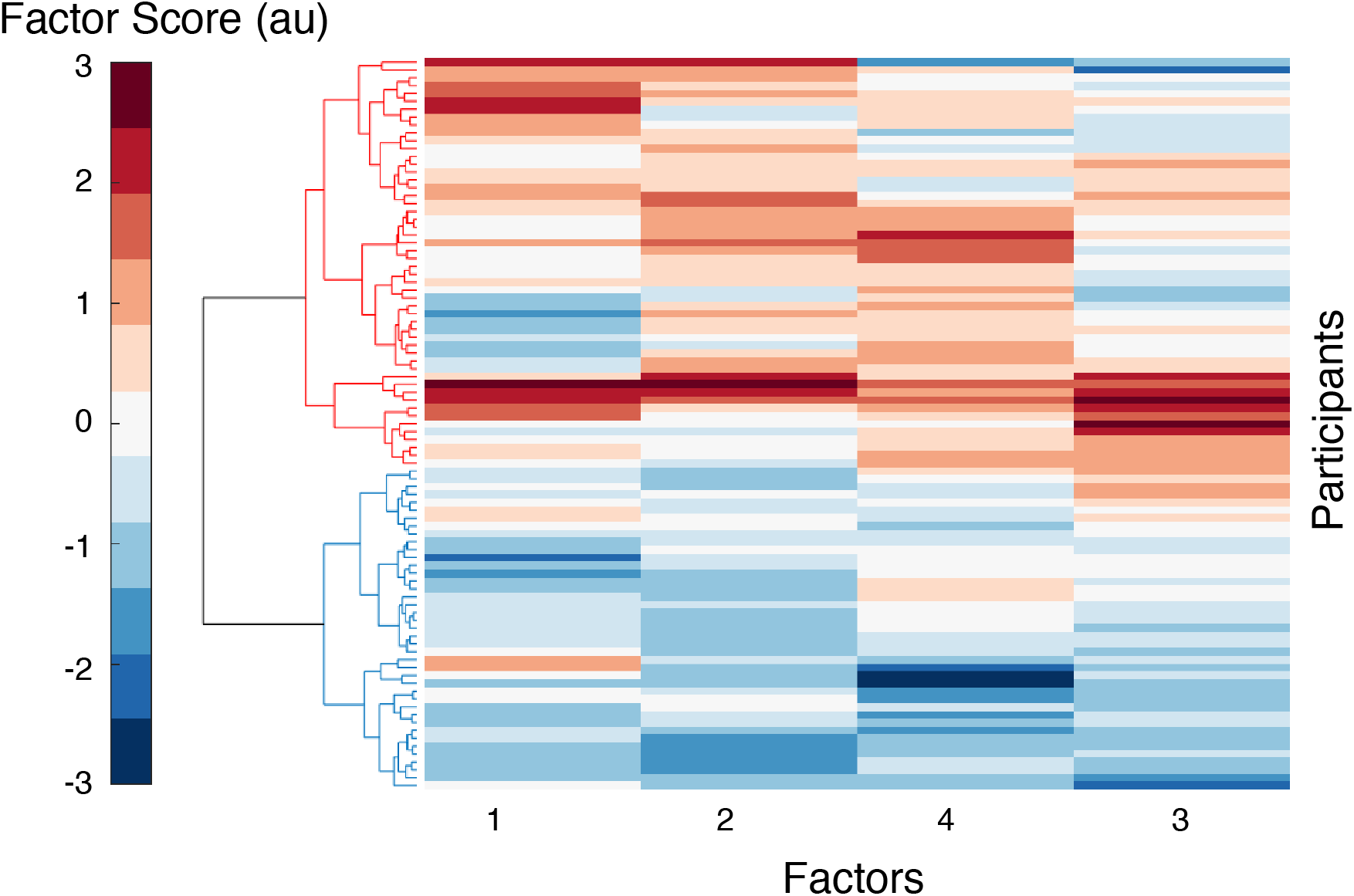
Clustergram: a matrix of each participants score across the 4 key factors identified by EFA. Factor score is measured in arbitrary units (au). Subjects form the y-axis, while each of the 4 factors is shown along the bottom. Factors 1 and 2 correspond to the mood and symptom burden, while Factors 3 and 4 correspond to the two of capability measures (physical and perceived, respectively). A dendrogram is displayed along the left side, highlighting the division of subjects into 2 clear groups.

**Figure 4.**
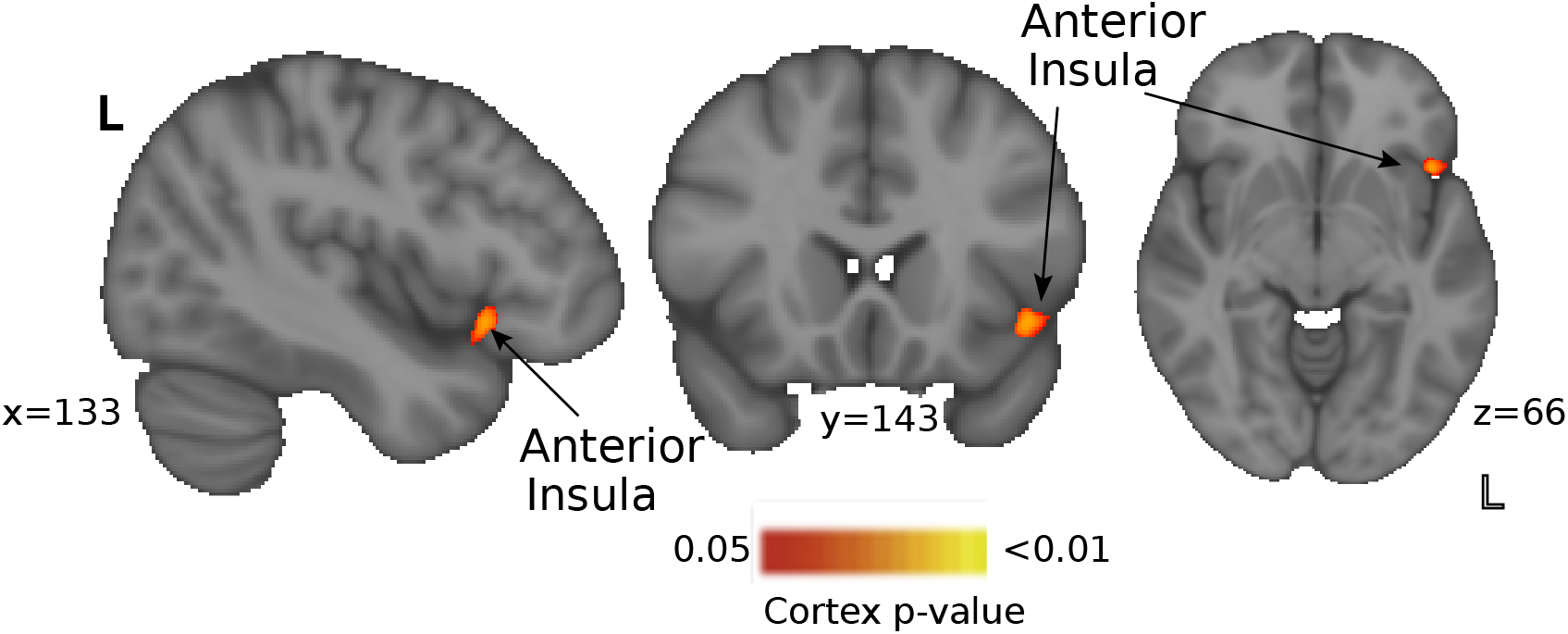
Blood Oxygen Level Dependent (BOLD) activity in response to breathlessness-related words compared to non-words between the two behavioural clusters of participants identified by the hierarchical cluster model (corresponding to high and low symptom load). Significant regional activity in the low symptom group than the high symptom group, was observed in anterior insula cortex with a non-parametric tfce p < 0.05.

#### Mean group differences – faces task

No difference in activity between groups in the condition of fearful versus happy faces survived. No significant difference was observed within the small ROI analysis of the amygdala. Additionally no significant difference was found in participant reaction times to happy (high load 758.57 ± 137.92 (ms), low load 782.09 ± 107.72 (ms)) or fearful faces (high load 742.71 ± 141.87 (ms), low load 773.70 ± 125.71 (ms)) (p>0.05).

#### 4. Stroop results

##### Group mean

**Table 4.** Reaction times of social and breathlessness-related anxious words compared to neutral words

|  | **Masked**<br>(mean ± SD (ms)) | **Unmasked**<br>(mean ± SD (ms)) |
| --- | --- | --- |
| Breathlessness Negative – Neutral | **22.5 ± 100.4** | **40.0 ± 114.9** |
| Social Negative - Neutral | **30.4 ± 109.5** | **2.6 ± 87.7** |

##### Group differences

**Table 5.** Reaction times of social and breathlessness-related anxious words compared to neutral words between the two groups

| **Masked** | **Group 1 (N=26)**<br>(mean ± SD (ms)) | **Group 2 (N=29)**<br>(mean ± SD (ms)) |
| --- | --- | --- |
| Breathlessness Negative – Neutral | **36.8 ± 98.6** | **25.0 ± 120.1** |
| Social Negative - Neutral | **62.1 ± 115.3** | **6.4 ± 132.7** |

| **Unmasked** | **Group 1 (N=26)**<br>(mean ± SD (ms)) | **Group 2 (N=29)**<br>(mean ± SD (ms)) |
| --- | --- | --- |
| Breathlessness Negative – Neutral | **50.1 ± 158.5** | **26.3 ± 113.0** |
| Social Negative - Neutral | **-13.3 ± 110.3** | **5.3 ± 81.5** |

In the masked condition, where emotional words were presented and then quickly followed by a symbol string mask, there was no significant difference between socially negative words and breathlessness-related words F(1,65)=0.09, p=0.76. However, in the unmasked condition, where emotional words were clearly visible and remained unmasked, there was a significant difference in reaction time in response to socially negative and breathlessness negative words F(1,65)=8.41, p<0.005. In this unmasked condition, participants responded significantly more slowly to breathlessness-related words.

Group identity was not found to be a significant predictor of difference in reaction time between socially negative words and breathlessness-related words in either the masked (F(1,53)=2.41, p=0.13) or unmasked conditions (F(1,53)=1.68, p=0.20)), nor were there any differences in reaction time in the neutral condition between the two groups (p>0.05).

Significant activity was observed in response to the breathlessness-related words compared to non-words, but not the fearful faces compared to happy faces. To address whether this could reflect specific breathlessness-related anxiety processing in this population, a direct contrast between the two conditions is shown in Supplementary Figure 8. This difference did not survive TFCE.

## Discussion

### Key findings

In this study we have stratified a group of people with COPD and found differences in their symptom perception, psychological profiles and brain activity in response to breathlessness-related word cues. Behaviourally, the two groups can be divided into high and low scorers on four key factors. These factors could loosely be considered to consist of one mood factor (Depression & Anxiety), one symptom burden factor (Vigilance, Catastrophising, Fatigue, St George – symptom, St George – impact, D12), and two capability factors – one physical (MSWT Distance, MSWT HR & St George – activity) and one anticipated (Avoidance – alone & accompanied & MRC). Importantly, no clear patho-physiological basis could be found to explain this group difference, based on the spirometry and oxygen saturation measures collected in this study.

Key differences in breathlessness-related brain activity were also found between the groups. These differences were observed within the anterior insula, a region that has previously been associated with the processing of bodily sensations and perceptions [3]. Interestingly, the differences in functional brain activity were only found in relation to specific breathlessness-related anxiety, with a more general anxiety paradigm utilising emotional faces revealing no significant differences between the groups, even when applying a region of interest approach within the amygdala, an area known to respond to threatening facial stimuli in highly anxious individuals. Indeed, across the whole group of participants, there was no significant difference in the brain response to fearful versus happy faces (Supplement Figure 7).

### A behavioural profile for chronic breathlessness

Accurate breathing perception is now thought to rely upon the carefully balanced integration of top down signals, which themselves arise from complex communications between different brain networks, and bottom up sensory information [6, 7]. Because of the multiple points of entry into this system, when the system is tipped out of balance, as can be the case in chronic breathlessness, aiming to pinpoint a single cause of disruption to the system is unlikely to reveal a universal mechanism in this individualised perception. If we accept that maladaptive-breathlessness can arise as a result of small shifts across a number of domains, including sensory, psychological and neurophysiological, a broad range of targeted measures are required to investigate these axes.

While each individual perceives breathlessness differently, based on this data, there may be subgroups of patients with similar underlying mechanistic disruptions to their symptom perception. An important first step towards subgroup stratification is to identify the best combination of measures capable of capturing the dimensions of the individual breathlessness experience. In the bourgeoning field of computational-psychiatry, big-data approaches are being applied to multimodal datasets. Where the conclusions of small sample size studies have been limited by cross population noise, the increased sensitivity afforded by machine learning techniques and larger sample sizes is now revealing neuro and behavioural phenotypes in conditions such as depression [25] and schizophrenia [26]. Such phenotypes may facilitate patient stratification, predict treatment outcomes and ultimately improve treatment success rates with personalised treatment plans. While our dataset cannot compare to these truly “big data” studies, where thousands of measures are available, the techniques employed here have been appropriately tailored from these studies for our purposes. In combination with detailed neuroimaging techniques, here we collected behavioural data from a population of people living with COPD. We sought to capitalise on this high-dimensionality to better understand breathlessness by uncovering data-trends ordinarily hidden by noise. While single measures may each offer a partial description of breathlessness, a more powerful approach is to combine relevant measures and draw out common factors. Variation across these factors may then link to neural networks or explain differences between individuals with objectively similar disease severity.

In this study, unsupervised machine learning algorithms indicated that 14/23 behavioural measures were important descriptors of the population. Collapsing across these measures revealed 4 key factors, which could loosely be considered to consist of one mood factor, a symptom burden factor and two capability factors – one physical and one anticipated. These measures paint an intuitive picture of the lived experience of breathlessness – what a person feels they *can* and *cannot* do, how their symptoms *impact* their lives and their general *mood*. Combining measures, where each addresses a small part of a larger, potentially biologically relevant factor, is perhaps the only way to access these concepts, and thus the neural activity that underlies the system.

The psychological impact of breathlessness-related cues was highlighted by the significant differences in reaction time observed in response to breathlessness-related words compared to socially negative words in an unmasked condition of the Stroop task. Participants responded more slowly to the breathlessness words, which could indicate that the salience of the breathlessness words were sufficiently attentionally demanding to absorb cognitive resources away from the task. This may reflect the greater attentional load of the breathlessness-related words compared to the socially negative related words. The finding was restricted to the unmasked condition suggesting that this may be a conscious process. This effect was not found to be group specific, although group difference was run on only a sub-sample of the participants, limiting direct comparisons. However, the absence of a group difference may also suggest that the impact of the breathlessness does not arise as a result of aberrant attentional processes.

Interestingly, physiological measures such as spirometry, pack-years and baseline oxygen saturation were not influential enough within the data variance to be included in the model, [1]. These findings lend further support to the strong impact of psychology and associated neural activity on the perception of breathlessness, and the importance of multimodal assessments.

### Stratifying patients based on behaviour

We then considered whether these four behavioural factors could be used to stratify the study population. The experience of chronic breathlessness is extremely heterogeneous, and a key question was whether evidence for subgroups could be revealed after the number of data dimensions were reduced down to just four core factors. Employing the technique of hierarchical cluster modelling, participants were grouped according to their scores on the four factors. Two clear groups were identified and validated via a scree plot, corresponding to high and low symptom load. Within each group, additional sub-groups could be observed, but participant numbers were such that these groups could not be easily interrogated further. Such groupings could of course be considered as existing as part of a broader distribution, where people fall onto a spectrum of symptoms. However, providing clear boundaries in this case enables us to initially consider the differences between people, which may relate to treatment options.

Following on from this, we were interested as to whether these behavioural groups corresponded to differences in brain activity patterns related to breathlessness. As part of the study, participants had completed two functional MRI scans – one a breathlessness-related word-cue task, to probe breathlessness specific expectation, and one a faces task, to probe general anxiety brain activity. Differences were observed across the two groups only for the word task. This finding mirrors those of *Rosenkranz et al*. [27], who noted that the insula was only activated by disease-related negative cues compared to generally-negative cues in a population of asthma patients.

Greater activity in response to word cues was observed within the anterior insula for the low symptom burden than high symptom burden. This area is part of the wider brain network thought to be involved with the affective perception of internal sensations and stimulus valuation [7]. Furthermore, the anterior insula has been linked to the processing of subjective feelings [3], and therefore, the greater activity within the network of Group 1 may be linked to a more coherent representation of internal sensations, leading to lower anxiety and general symptom burden. Indeed, the specificity of this group difference to breathlessness supports the theory that this network is specific and potentially related to sensory interpretations. However, further studies utilising measures that directly measure symptoms against breathlessness perceptions would be required to investigate this hypothesis.

To tease apart the mechanisms of breathlessness, the study of perception is beginning to take on lessons from Bayesian theories to frame itself in terms of priors, predictions and likelihoods. From this perspective, recent work suggests a complex interplay between descending top down control from stimulus valuation areas such as the anterior insula, anterior cingulate cortex, orbitofrontal cortex and ventromedial prefrontal area, and ascending sensory signals received by areas such as periaqueductal gray, thalamus and posterior insula [9] [5]. Such models could explain why pulmonary rehabilitation is such an effective treatment for breathlessness, despite the lack of any effect on lung function Furthermore, reductions in breathlessness anxiety resulting from pulmonary rehabilitation have been found to correlate with activity in the insula and anterior cingulate cortex; key areas identified for stimulus evaluation [9].

### Further considerations and limitations

Although the techniques employed here draw upon the big data techniques of machine learning, the acceptable ratio between variables and participants must be carefully considered when scaling down these techniques. Factor analysis requires between 2 and 5 times the number of samples per variable tested and ideally more than 100 observations. With further subjects it may also be possible to gain sufficient statistical power to further probe further sub-groups, such as those visible within Figure 3. Another important caveat of cluster methods is that they examine shared variance, and so we must remain aware that any of the measures not included in this model could be highly relevant – but not share any common features with the other measures.

Incorporating psychological variables into models of breathlessness to investigate the influence of mood and previous experience enriches our ability to explain potential influences towards breathlessness perceptions. However, in this cross-sectional work, the direction of causality is impossible to infer - are people with low mood more susceptible to breathlessness anxiety, or do they become low in mood as a result of their greater symptom burden? Longitudinal population studies would thus need to be employed to understand any predominating direction within this relationship.

## Conclusion

Breathlessness perception in people living with COPD is highly heterogeneous and frequently does not match their medically diagnosed disease status. Given the relatively high prevalence of COPD within the general population addressing this discrepancy has the potential to meaningfully impact hundreds of thousands of lives. The results from the current study thus move us towards a more comprehensive understanding of the contributing components influencing breathlessness perception. We have identified both perceptual and breathlessness-related brain activity differences within two subgroups of our COPD patients, demonstrating distinctive neuro-psychological profiles between those with high and low symptom loads. While further work is required to elucidate more nuanced subgroups and components relating to individualised breathlessness perception, we have demonstrated that incorporating behavioural and neuroimaging measures into enriched models can offer new perspectives on breathlessness, and move us towards the goal of tailoring treatment programs for the individual and their lived experience of breathlessness.

## Data Availability

Data available by contacting authors subject to data sharing policies of the Nuffield Dept Clinical Neurosciences

## Acknowledgements

This work was supported by the Dunhill Medical Trust (Grant R333/0214) and the National Institute for Health Research Biomedical Research Centre (Grant RCF18/002) based at Oxford University Hospitals NHS Foundation Trust and The University of Oxford.

CJH is supported by the National Institute for Health Research Biomedical Research Centre based at Oxford Health NHS Foundation Trust and The University of Oxford, and by the UK Medical Research Council.

## Supplementary Material

### Disclosure

This body of work was collected as part of a larger project investigating the effect of D-cycloserine (a partial NMDA receptor agonist, with target sites that include the amygdala) on outcome measures of pulmonary rehabilitation. This population forms the baseline component of this study and full results of the trial will be published separately and at a later date.

### Recruitment

**Figure S1.**
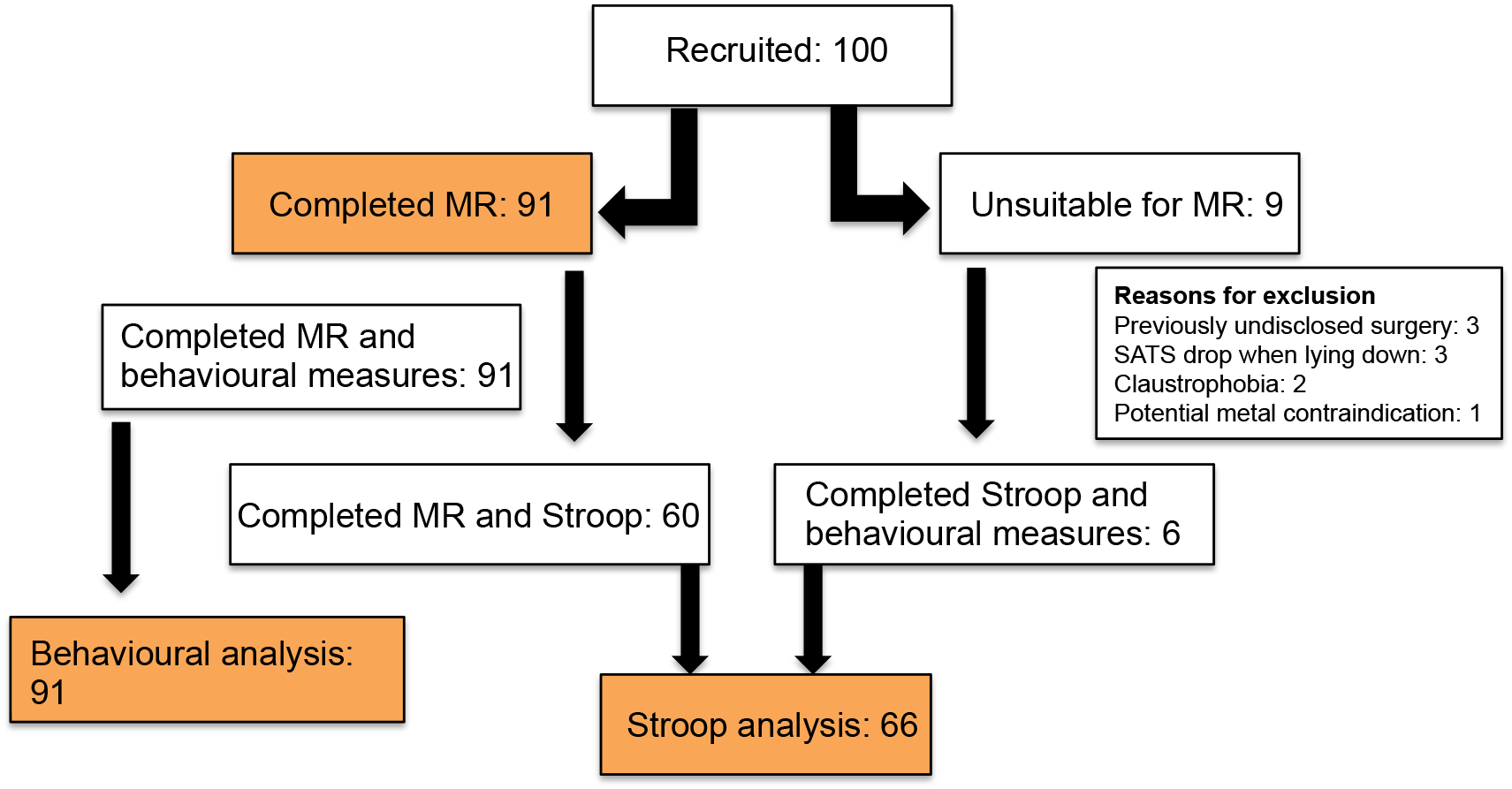
Consort diagram illustrating participant progression through the study. Key analysis points, shown in orange, were MR analysis (91 participants), behavioural analysis (91 participants) and Stroop analysis (66 participants).

### Participants

Study inclusion criteria included a diagnosis of COPD and admittance to pulmonary rehabilitation. Exclusion criteria included inadequate understanding of verbal and written English, significant cardiac, psychiatric (including depression under tertiary care) or metabolic disease (including insulin controlled diabetes), stroke, contraindications to d-cycloserine (including alcoholism), epilepsy, claustrophobia, regular therapy with opioid analgesics or oxygen therapy. Of the 100 participants recruited, 91 completed the MRI component of the study. Reasons for non-compliance with the MRI component of the study included claustrophobia, unchecked surgical implants, concern regarding potential previous metal injury and inability to lie supine for 40 minutes.

### Behavioural Measures

#### Questionnaire Measures

##### Dyspnoea-12 (D12) Questionnaire

This is a 12-item questionnaire designed to measure the severity of breathlessness and has been validated for use in patients with respiratory disease [1].

##### Centre for Epidemiologic Studies Depression Scale (CES-D)

Depressive symptoms are commonly observed in patients with respiratory disease. This brief questionnaire consists of 20 items investigates the symptoms of depression across a number of factors [2].

##### Trait Anxiety Inventory (TRAIT)

This questionnaire assesses participant’s general level of anxiety in particular scenarios via 20 questions asking “how anxious you generally feel” [3].

##### Fatigue Severity Scale

This 9-point questionnaire quantifies patient fatigue, which is well documented in its association with COPD [4].

##### St George’s Respiratory Questionnaire (SGRQ)

There are 50 questions in this questionnaire, which has been developed and validated for use in COPD and asthma. The questions measure the impact of overall health, daily life and well-being [5].

##### Medical Research Council (MRC) breathlessness scale

The MRC scale quantifies perceived difficulty due to respiratory restrictions on a scale of 1 to 5 [6].

##### Mobility Inventory (MI)

This questionnaire collects data regarding the extent to which a participant avoids certain situations, either alone or accompanied (21-items in each category) [7].

##### Pittsburgh Sleep Quality Index

The 17 items of this questionnaire examine self-reported quality, duration and regularity of sleep [8].

##### Anxiety Sensitivity Index Questionnaire (ASI)

This 16-item questionnaire probes the somatic, cognitive and social facets of anxiety and its harmful consequences. The ASI has been validated in distinguishing between clinical and non-clinical populations and demonstrates high test-retest reliability [9].

##### Catastrophic Thinking Scale in Asthma

This 13-point questionnaire was modified for this study by substituting the word “asthma” for “breathlessness” in order to measure catastrophic thinking [10] [11].

##### Pain Awareness and Vigilance Scale

This questionnaire was modified by substituting the word “breathlessness” for the word “pain”. The 16-point scale measures how much a participant focuses their attention onto their breathlessness [12] [11].

### Emotional Stroop

Biased cognitive processing relating to perceived threatening stimuli may relate to the phenotype of breathlessness in COPD. Attentional bias has been observed in panic disorder [18], as well as in anxiety and depression, [19], which are highly co-morbid with chronic breathlessness. Indeed, in their 2019 paper Sucec and colleagues suggest that the threat of breathlessness may affect attentional processing by absorbing the available resources [20]. We assessed whether these findings could be replicated within the COPD population using an emotional Stroop task. Emotional words were selected on the basis of patient interactions. The valence of these words was then tested in a population of 14 control participants and 17 COPD patients. From this pool of words, 12 breathlessness-negative words were selected and were combined with the 12 socially-negative and 12 neutral words from [18]. Due to technical issues with the emotional Stroop task (data loss due to computer malfunction), data was collected from a subgroup of 66 of the 100 participants. Analysis for this task was kept separate from the behavioural correlation analysis due to the large number of missing data points. Of the 66 participants who completed the Stroop task, complete MRI data was available for 55 participants.

Stimuli were programmed using e-prime (Psychology Software Tools, 2002) and consisted of 12 neutral words (e.g. Vase), 12 social-threat words (e.g. Stupid) and 12 breathlessness-threat words (e.g. Choking) presented at font size 24. Stimuli were presented in six experimental blocks; one for each word type (neutral, social-threat and breathlessness-threat) in both masked and unmasked conditions. In the masked condition, target stimuli were presented and then replaced by a symbol string matched for font size, colour and length after 17ms. Block order was counterbalanced across participants. Participants were instructed to name the colour of the word (red, blue or green) as quickly and accurately as possible, with the word remaining visible to the participant until a response was made. Participant responses were recorded using a microphone, which was connected to a laptop via a serial response box. The serial response box converted the initiation of a vocalisation into a temporal signal, enabling reaction time to be established, while the experimenter recorded accuracy with a button press.

### Physiological Measures

A trained respiratory nurse collected spirometry measures of FEV1 and FVC using Association for Respiratory Technology and Physiology standards. Participants performed two incremental shuttle walk tests (MSWT), and heart rate and oxygen saturations (SpO_2_) were measured immediately before the MSWT and subsequently every minute until 10 minutes post-exercise (or until participants returned to their baseline state) using a fingertip pulse oximeter (Go_2_; Nonin Medical Inc). Before and after the IWST participants also rated their breathlessness on a modified Borg scale [21].

### Imaging Measures

#### Word Task

Learned associations can be employed to indirectly probe related brain networks and the cognitive dynamics of emotional states such as anxiety and fear. A relevant example could include a healthy person needing to walk up several flights of stairs, who may think nothing of the experience, whereas for a person with a lung condition the stairs may be associated with memories of fearful breathlessness. Given sufficient repeated threatening exposures, the thought alone of climbing stairs may trigger the networks sub-serving the corresponding anxiety response. In such cases, the presentation of a cue in the form of a word has been shown sufficient to activate brain networks related to the action or emotional state associated with this cue. This task was developed and published by Herigstad and colleagues in 2016 for use in the COPD population [13]. Word cues were developed in three key stages; firstly in collaboration with respiratory practitioners, academics and physiotherapists, a set of 30 word cues associated with breathlessness were created. Next, these cues were provided to patients with COPD alongside a VAS rating scale, allowing patients to rate how breathless and anxious the situations identified by the cues would make them feel. Following adjustments based on participant feedback, the word cues were then computerised and tested in a larger population of COPD patients [13]. Further validation was carried out in the fMRI environment and by for clinical sensitivity with comparisons between changes in key questionnaire measures and word-cue rating. In the first scan, participants were shown a set of breathlessness-related word cues in a pseudo-randomised order. During the task, participants were presented with a word cue in white text on a black background for 7 seconds. Participants were then asked, “how breathless would this make you feel” (wB) and “how anxious would this make you feel” (wA). To each question participants responded within a 7 second window using a button box and visual analogue scale (VAS). The response marker always initially appeared at the centre of the scale, with the anchors “Not at all” and “Very much” at either end. Before the scan session, participants were given the opportunity to practice using the button box with a set of test words. A control condition, used as a baseline measure of activity in response to the presentation of a visual stimulus was presented 4 times over the course of the scan, consisting of a string of “XX” with fixed length of 15 characters, and each time was presented for 7 seconds. No rating period followed these control blocks. Subtracting activity in response to the control condition (i.e. simply the response to any visual stimulation) from conditions of interest enabled us to examine stimulus-specific activity.

#### Faces Task

Emotional facial expressions are widely recognised to activate the same neural pathways as the behavioural emotion conveyed by the expression itself [14]. Fearful facial expressions, for example, have been shown to correspond to activity within the amygdala, a region known to modulate fear processing [15]. The speed and accuracy of task completion, which in this instance was the recognition of facial gender, under different emotional conditions can be used probe whether response bias exists. This draws upon work suggesting that the threat of breathlessness may absorb cognitive resources and affect the processing of new information via attentional bias [16, 17]. Biased attentional processes, either towards or away from potentially threatening situations, may be mirrored by activity patterns within threat and fear brain networks.

### MRI Acquisition

Prior to each MRI session participants were screened for standard MRI contraindications including metal in or about their person, epilepsy and claustrophobia.

#### Sequence Parameters

T1 sequence parameters: TR, 2040ms; TE, 4.68ms; voxel size, 1 × 1 × 1 mm; FOV, 200mm; flip angle, 8°; inversion time, 900ms; bandwidth 130 Hz/Px).

T2*-weighted (functional) sequence parameters: TR, 3000ms; TE 30ms; voxel size 3 × 3 × 3 mm; FOV, 192mm; flip angle 87°; echo spacing 0.49ms.

Functional scan durations: word-task - 215 volumes, 10 minutes and 27 seconds duration and faces task - 168 volumes, 8 minutes and 24 seconds duration.

Field map sequence parameters: TR, 488ms; TE1, 5.19ms; TE2, 7.65ms; flip angle 60°; voxel size, 3.5 × 3.5 × 3.5 mm.

### Analysis

#### Cluster Analysis

When calculating the full correlation matrices for the hierarchical clustergrams, scoring was reversed for the following measures: MSWT Distance, MSWT starting oxygen saturation, MSWT heart rate change and spirometry. This ensured that higher scores were associated with worse symptomatology across all measures.

Hierarchical cluster models reorder variables based on their correlation strengths so that groups of related measures sit closer to each other than non-related measures.

This allows natural relationships to be easily visualised. The modeling process formalises not only the relationship between pairs of variables, but also the manner by which shared variance can be described as part of larger, related clusters. The clustering algorithm initially considers pairs of variables in terms of their similarity or “distance” (in arbitrary units). Linked pairs are then incorporated into larger clusters with the goal of minimizing a cost function (distance to be bridged), a process that can be thought of as minimizing the dissimilarity within clusters. As pairs become clusters, a cluster tree or dendrogram is created. The distance between neighbouring branches indicates the relative similarity of two measures, while advancing up the hierarchical cluster tree moves further away in terms of link distance, and therefore similarity.

Hierarchical models are useful as a descriptive tool for examining and visualising the structure of the dataset as a whole. However, they do not provide information as to the significance of any given cluster of behavioural measures. In contrast, exploratory factor analysis (EFA), which falls under the umbrella of structural equation modeling, can be used to formalise the relationships observed in the hierarchical models. This allows the researcher to establish the presence of underlying shared constructs via a number of fit statistics without applying a preconceived structure on the result.

For the EFA analysis, the smallest number of uncorrelated clusters that maximally explain the variance of the dataset was estimated. In this instance, parallel analysis with oblique rotation was employed to calculate this value and the results were visualised using a scree plot (Supplementary Figure 3). In a second step, the number of variables to be retained within the model was determined. A maximum likelihood estimation approach was applied, where variables that did not load significantly onto a particular factor, or demonstrated significant cross loading (i.e. loaded onto more than one factor) were excluded from further testing. Finally, the model statistics were interrogated to formalize the shared variance across latent factors and the extent to which each variable contributed to its factor as a whole. The smallest number of factors that significantly explained the variance across the dataset was then accepted as the model of best fit. Model selection criteria included loading variables above 0.4 with no cross loading or freestanding variables, and significant X^2^/df ratio with Tucker-Lewis Index (TL-index) close to 1 and RMSEA < 0.06. Models were fit using Lavaan version 0.6-1 [22] in R version 3.2.1 (R Core Team).

#### Imaging Analysis

##### MRI Preprocessing

The data were corrected for movement using MCFLIRT (Motion correction using FMRIB’s Linear Image Registration Tool [23]). Non-brain structures were removed using BET (Brain Extraction Tool [24]). Spatial smoothing was carried out using a full-width-half-maximum Gaussian kernel of 5mm, while high-pass temporal filtering (Gaussian-weighted least squares straight line fitting; 90 s) removed low frequency noise and slow-drift.

Distortion correct of EPI data was carried out using a combination of FUGUE (FMRIB’s Utility for Geometrically Unwarping EPI’s [25, 26] and BBR (Boundary Based Registration; part of the FMR Expert Analysis Tool, FEAT version 6.0 [27]).

Data denoising was carried out as follows: Before the first level analysis, each functional scan was decomposed into maximally independent components using FMRIB’s MELODIC tool (Multivariate Exploratory Linear Optimised Decomposition into Independent Components). “Noise” components were identified by FIX (FMRIB’s auto-classification tool, [28, 29]) using the WhII.Standard.RData [30] trained classifier with aggressive clean up option. A Principle Component Analysis (PCA) was run on the FIX identified components to retrain 99% of the variance. Separately, the cardiac and respiratory related physiological signals (recorded via a pulse oximeter and a respiratory bellows) were transformed into a series of regressors, (three cardiac and four respiratory harmonics) as well as an interaction term and a measure of respiratory volume per unit of time (RVT), using FSL’s physiological noise modeling tool (PNM). The signal associated with these waveforms (modeled using retrospective image correction (RETROICOR) [31, 32]) was then used to form voxelwise noise regressors.

The confounds identified by FSL’s FIX and PNM tools, along with sources of noise arising from motion, were then combined into a single model. This single noise model approach builds upon the technique outlined by [33]; and fully detailed by [34]. In these preceding works we employed a step-wise technique whereby physiological noise (identified by PNM) and FIX-identified noise were each removed from the data in separate steps prior to data entry into the lower level model. In the new cleanup pipeline, a single text file containing time-course information relating to FIX identified noise components along with white matter or CSF related noise was included as additional confound EV’s within the lower level model, while the PNM-identified noise was entered into the model as a standard voxel-wise confound list. In this updated de-noising pipeline, confounds identified above are added to model at the stage of first-level analysis and thus the functional dataset can be corrected for sources of noise arising from motion, scanner and cerebro-spinal fluid artefacts, cardiac, and respiratory noise in a single step, rather than three.

##### Image Registration

The functional scans were registered in a two-step process to the MNI152 (1×1×1 mm) standard space brain template. Firstly, each subject’s EPI was registered to their associated T1-weighted structural image using BBR (6 DOF) with nonlinear field map distortion correction [27]. In the second step the subject’s structural image was registered to 1mm standard space via an affine transformation followed by nonlinear registration (using FNIRT: FMRIB’s Non-linear Registration Tool [35]).

##### Statistical Thresholds

This study was originally powered to test for significance at z>2.3, p<0.05, prior to the adoption of the now standard cluster correction threshold of z>3.1 or threshold free cluster enhancement. Therefore, we have additionally provided a supplementary demonstration (Supplementary Figure 6) of the significant activity at this reduced threshold.

### Results

The value of combining EFA with a hierarchical cluster model can be visualised by comparing Supplementary Figure 2 with Figure 3. In Supplementary Figure 2 all available behavioural measures have been included in a hierarchical cluster model. As a result, while a colour gradient top-bottom (red to blue respectively) is roughly visible, no clear structure can be seen in the dendrogram shown left of the figure. In contrast, once the number of dimensions have been reduced, as shown in Figure 3, clear groups can be observed in the dendrogram – shown left of the central correlation matrix

Further characterization of the fMRI results can be found in supplementary Figure 5, where the response to breathlessness-related words compared to non-words in all participants (A), and in the high and low symptom load groups is shown. The contrast of activity in response to breathlessness-related words compared to non-words between the two behavioural clusters of participants is shown in Supplementary Figure 6. Supplementary Figure 7 shows the activation for fearful and happy faces across all participants (no significant difference between groups was found).

In a direct comparison of the two tasks across the groups of participants (high and low symptom load), significant activity was observed within the anterior insula, middle frontal gyrus and post-central gyrus in the low symptom load group to the breathlessness-related fear cues (Supplement Figure 8). This difference did not survive the correction with more rigorous thresholds now expected of neuroimaging studies.

**Figure S2.**
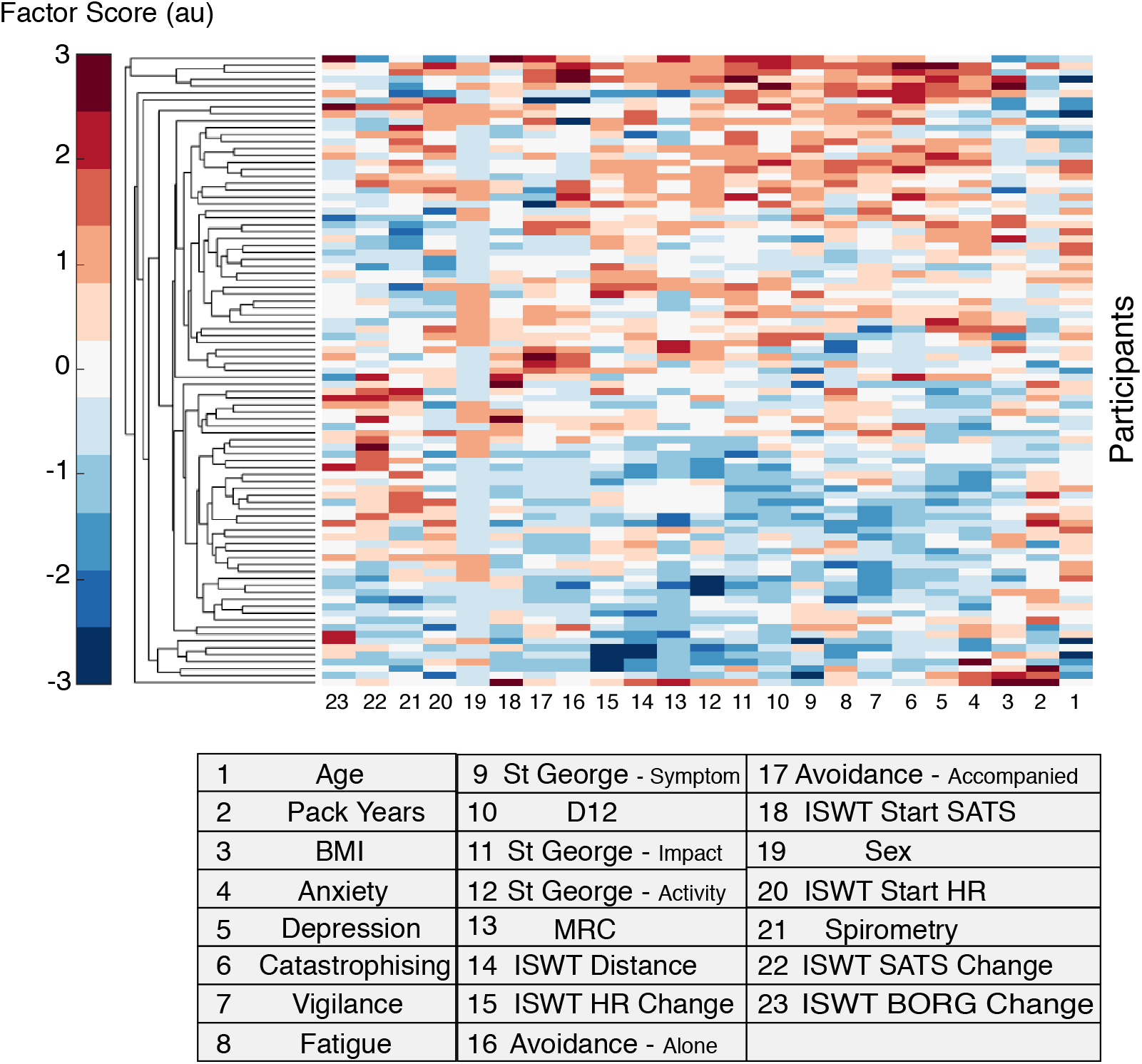
Clustergram: a matrix of each participants score across the all measures (shown within the table). Factor score is measured in arbitrary units (au). Participants form the y-axis, while each variable is shown along the bottom. A dendrogram is displayed along the left side with no clear subject grouping.

**Figure S3.**
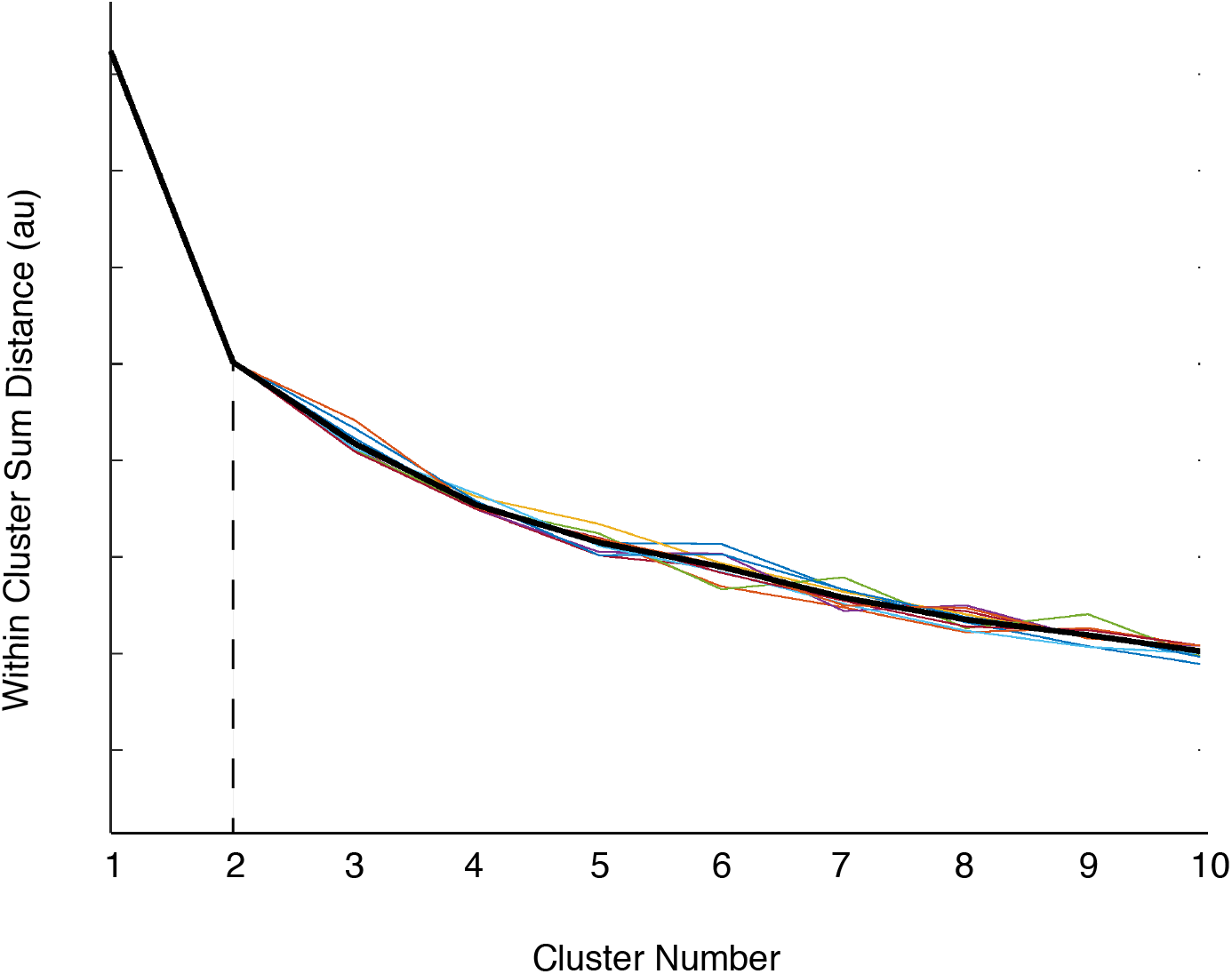
Scree plot - where within cluster distance (y-axis, au –arbitrary units) is plotted as a function of cluster number (x-axis). The point at which the addition of further clusters no longer significantly explains more of the dataset variance can be visualized as an “elbow” in the plot. Each of the coloured lines represents a single trial. The thicker black line represents the average of all trials (N=10). The dashed line highlights the elbow point of the graph, in this instance at 2 clusters.

**Figure S4.**
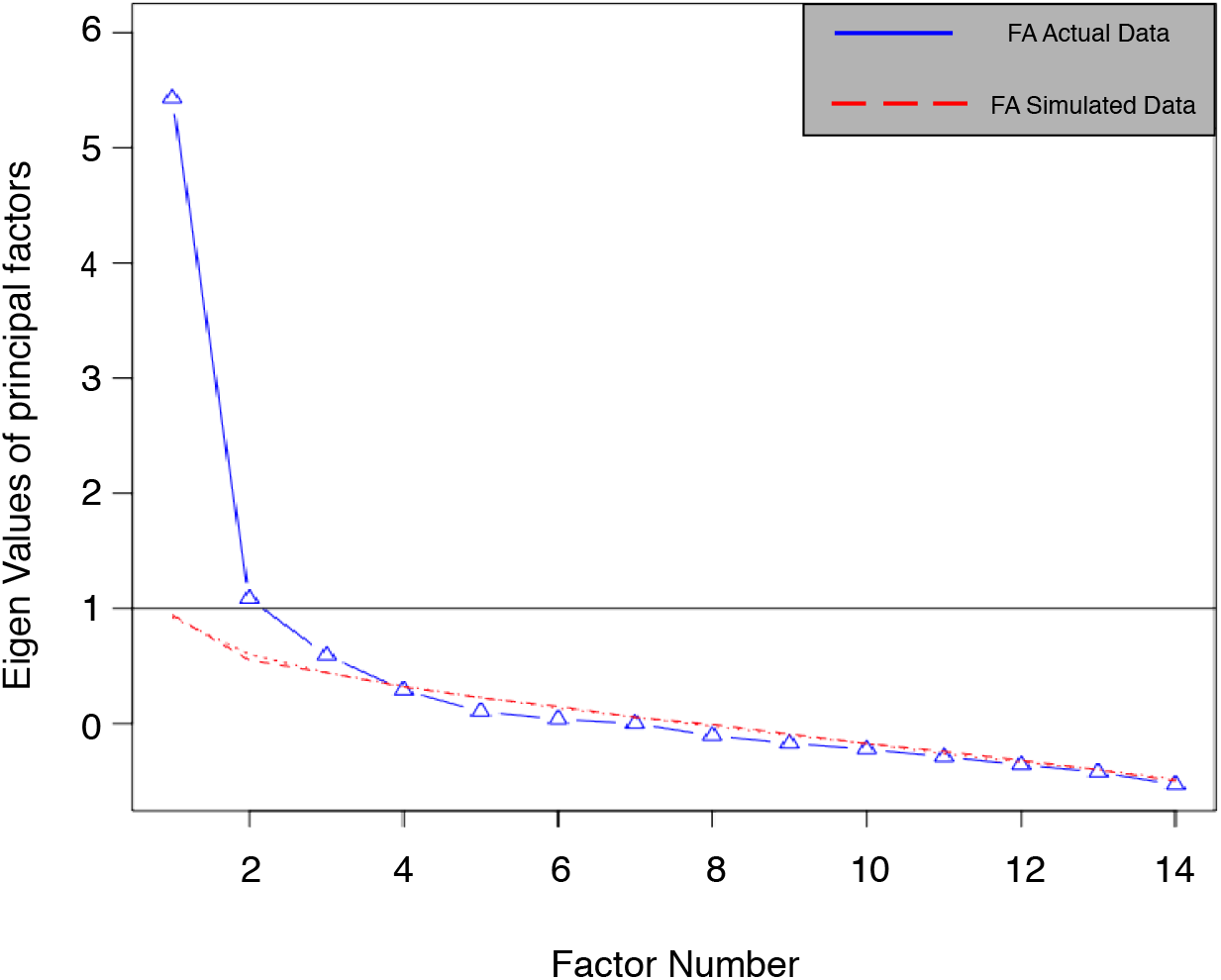
Scree plot – Created using parallel analysis, where the eigenvalues of the real data (blue solid line) are shown for each factor extracted compared to extracting the same number of factors from a similarly sized random dataset (red dotted line). Eigenvalues are shown on the y-axis, while factor number is plotted on the x-axis. The point at which the Eigenvalues of the real dataset are no longer greater than that drawn from the random dataset indicates the most distinct number of factors.

**Figure S5.**
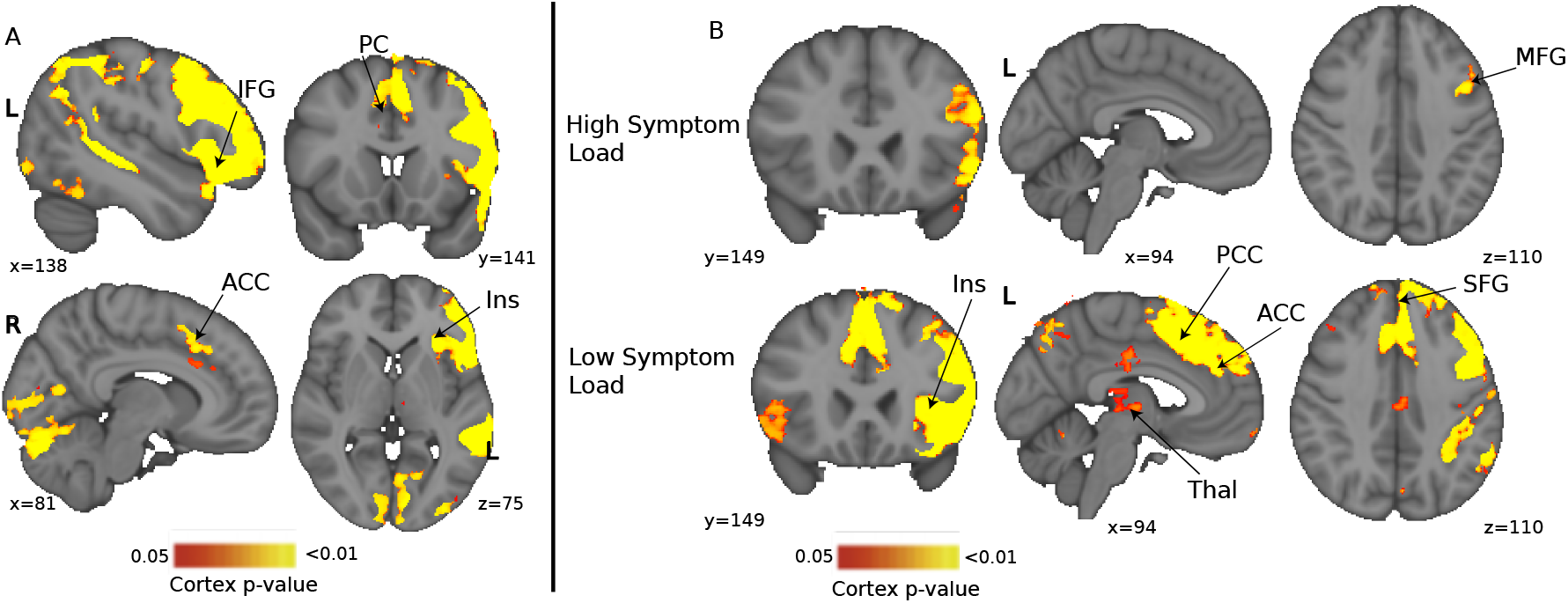
Blood Oxygen Level Dependent (BOLD) activity in response to breathlessness related words compared to non-words in (A) across all participants, and (B) for high and low symptom load groups separately. Significant regions, active in the group mean, including Inferior Frontal Gyrus (IFG), Posterior Cingulate (PC), Anterior Cingulate Cortex (ACC) and Insula (Ins) are displayed with a non-parametric tfce p < 0.05. Inferior frontal and middle front gyri demonstrated significant activity in the high symptom group. In the low symptom group, ParaCingulate Cortex (PCC), ACC, insula, Thalamus (Thal) and Superior Frontal Gyrus (SFG) all demonstrated significant activity. For both high and low symptom groups, significant regions are displayed with a non-parametric tfce p < 0.05.

**Figure S6.**
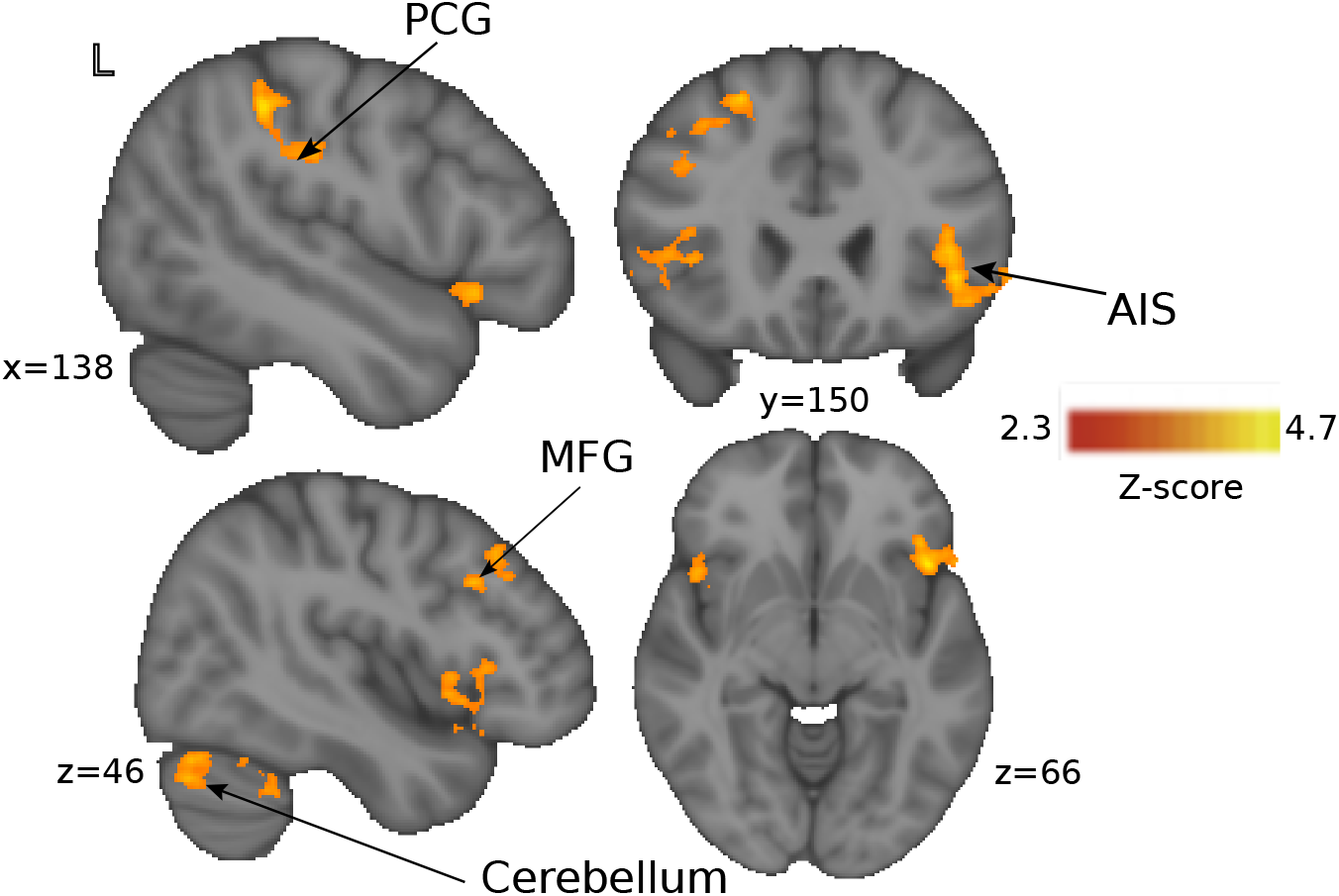
Blood Oxygen Level Dependent (BOLD) activity in response to breathlessness related words compared to non-words between the two behavioural clusters of participants identified by the hierarchical cluster model (corresponding to high and low symptom load). Significant regions demonstrating greater activity in the low symptom group than the high symptom group include Anterior Insula (AIS), Medial Frontal Gyrus (MFG), Post Central Gyrus (PCG) and Cerebellum. Significance is expressed at Z>2.3, p<0.05.

**Figure S7.**
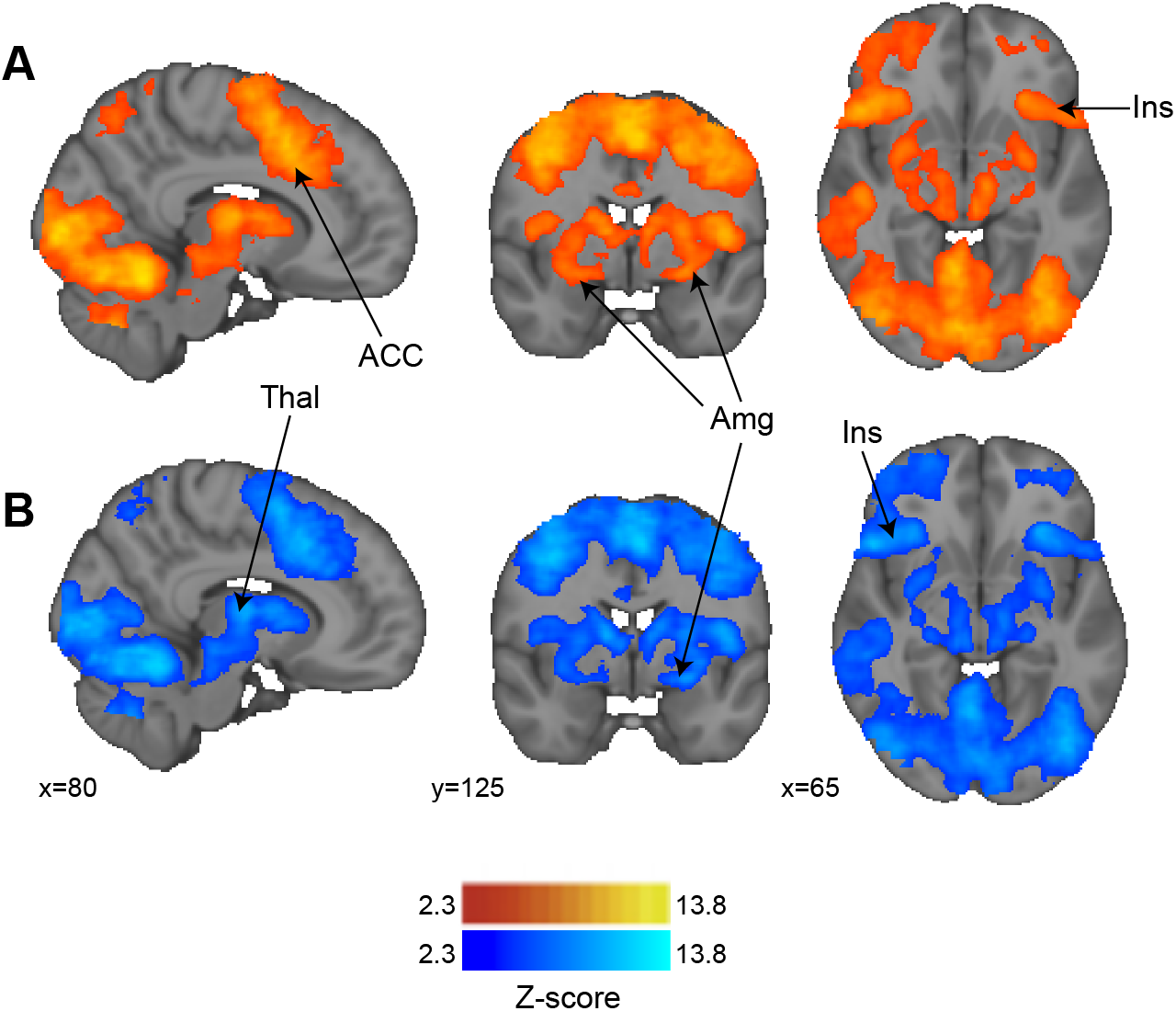
Blood Oxygen Level Dependent (BOLD) activity in response to happy faces (A) and fearful faces (B) across all participants. Significant (Z>2.3, p<0.05) activity was observed within the anterior cingulate cortex (ACC), amygdala (Amg), insula (Ins) and temporal and prefrontal thalamus (Thal) in response to happy and fearful faces. The contrast of the two conditions did not reveal any significant differences.

**Figure S8.**
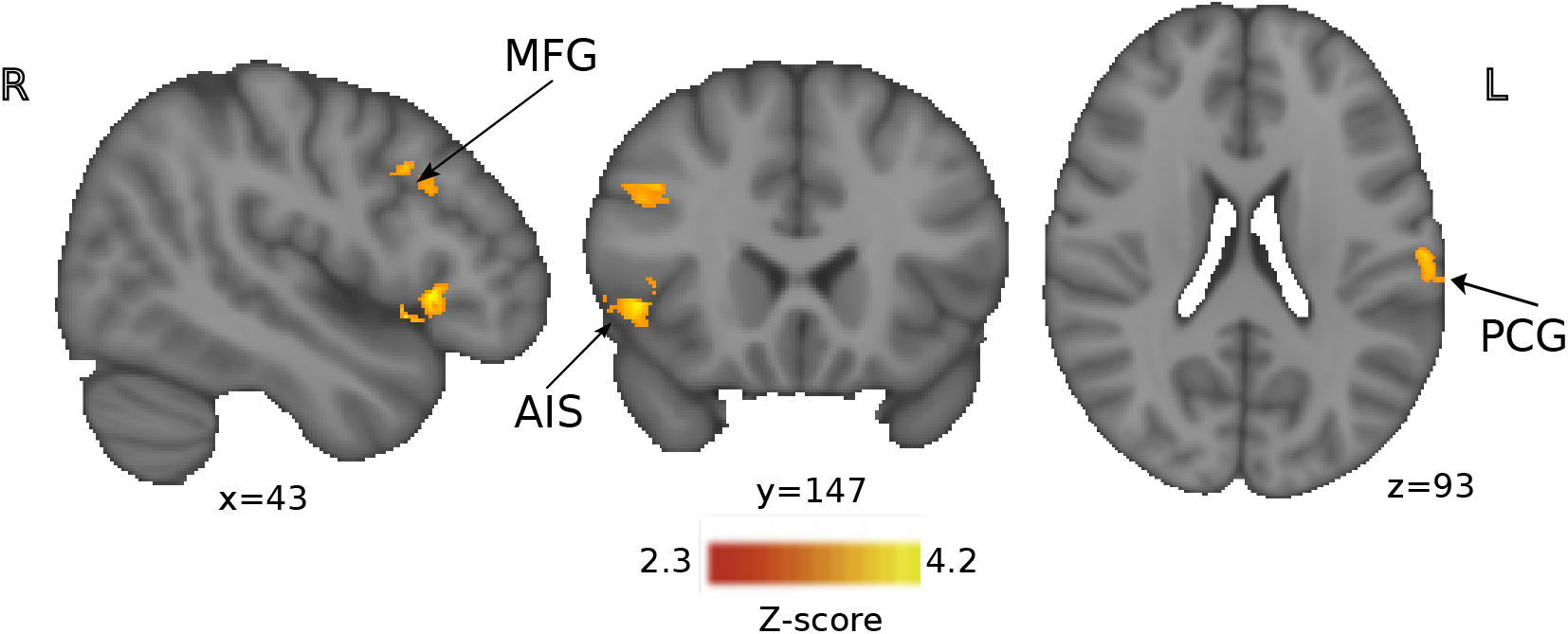
Blood Oxygen Level Dependent (BOLD) activity in response to breathlessness related words (greater than non-words) compared to fearful faces (greater than happy faces) between the two behavioural clusters of participants identified by the hierarchical cluster model (corresponding to high and low symptom load). Significant activity (Z>2.3, p<0.05) was observed within the Anterior Insula (AIS), Medial Frontal Gyrus (MFG), Post Central Gyrus (PCG) in the low symptom load greater than the high symp tom load.

